# Slide-free surface histology enables rapid colonic polyp interpretation across specialties and foundation AI

**DOI:** 10.1101/2025.06.11.25329397

**Authors:** Alexander Si Kai Yong, Nazihah Husna, Ko Hui Tan, Gaurav Manek, Rachel Rui Yi Sim, Rachel May Ling Loi, Oswald Zhao Jian Lee, Sarah Shuyun Tang, Gwyneth Shook Ting Soon, Dedrick Kok Hong Chan, Kaicheng Liang

## Abstract

Colonoscopy is a mainstay of colorectal cancer screening and has helped to lower cancer incidence and mortality. The resection of polyps during colonoscopy is critical for tissue diagnosis and prevention of colorectal cancer, albeit resulting in increased resource requirements and expense. Discarding resected benign polyps without sending for histopathological processing and confirmatory diagnosis, known as the ‘resect and discard’ strategy, could enhance efficiency but is not commonly practiced due to endoscopists’ predominant preference for pathological confirmation. The inaccessibility of histopathology from unprocessed resected tissue hampers endoscopic decisions. We show that intraprocedural fibre-optic microscopy with ultraviolet-C surface excitation (FUSE) of polyps post-resection enables rapid diagnosis, potentially complementing endoscopic interpretation and incorporating pathologist oversight. In a clinical study of 28 patients, slide-free FUSE microscopy of freshly resected polyps yielded mucosal views that greatly magnified the surface patterns observed on endoscopy and revealed previously unavailable histopathological signatures. We term this new cross-specialty readout ‘surface histology’. In blinded interpretations of 42 polyps (19 training, 23 reading) by endoscopists and pathologists of varying experience, surface histology differentiated normal/benign, low-grade dysplasia, and high-grade dysplasia and cancer, with 100% performance in classifying high/low risk. This FUSE dataset was also successfully interpreted by foundation AI models pretrained on histopathology slides, illustrating a new potential for these models to not only expedite conventional pathology tasks but also autonomously provide instant expert feedback during procedures that typically lack pathologists. Surface histology readouts during colonoscopy promise to empower endoscopist decisions and broadly enhance confidence and participation in ‘resect and discard’.

**One Sentence Summary:** Rapid microscopy of resected polyps during colonoscopy yielded accurate diagnoses, promising to enhance colorectal screening.

## INTRODUCTION

Colorectal cancer is the third most common cancer and second leading cause of cancer death worldwide(*1*). Colonoscopy is a mainstay tool in colorectal cancer screening, with substantial improvements in survival rates over recent decades(*2*). However, the cost of the procedure remains prohibitively high, largely due to the need for both trained endoscopists and reliable histopathology facilities and expertise. One key driver of cost is the handling of colonic polyps, anomalous tissue growths that are resected during the procedure and incur expensive processing and histological interpretation. A substantial proportion of resected polyps are benign and thus of low diagnostic value, but are nevertheless commonly sent for histology. One approach towards enhanced efficiency is ‘resect and discard’ – to discard resected polyps with a benign endoscopic appearance without sending for pathological diagnosis. However, despite many studies affirming endoscopic diagnosis including a large successful multicentre trial(*3*), ‘resect and discard’ is still rarely performed even in the countries that led these efforts. Consequently, progress in reforming existing practice has been exceedingly slow.

One possible explanation for the low adoption rate of ‘resect and discard’ is that most endoscopists remain uncertain from endoscopic interpretation alone, and still prefer pathologist confirmation. If so, the ‘resect and discard’ paradigm may be significantly strengthened with pathologist signoff on real-time findings from endoscopy, allowing a rebalancing of risk more faithful to classical practice. However, while endoscopists and pathologists are both experts in image interpretation, conventions of image acquisition and appearance are vastly different. Crucially, pathology slides require arduous tissue preparation and have stringent requirements on microscopic image quality and interpretation that are typically incompatible with real-time feedback. Pathologists may provide rapid feedback on tissue via frozen sections in certain surgeries where resection margins are critical(*4*), but virtually never in endoscopic procedures.

Slide-free histology is a new approach to real-time tissue diagnosis, using advanced microscopes capable of imaging fresh unprocessed (unsectioned) samples to derive histological readouts, as an alternative to frozen sections. Successful laser microscopes for imaging of unsectioned tissue to guide surgery include multiphoton(*5*, *6*), Raman(*7*), or light sheet(*8*). However, in the unique context of the endoscopic suite, low-cost and laser-free modalities are preferred for ease of adoption, compactness and reliability. Widefield microscopy using ultraviolet excitation is a recent laser-free approach(*9–12*) that has been successful in imaging of thick fresh clinical samples despite its relative simplicity in optical implementation.

Here we present a successful intraprocedural deployment of a state-of-the-art ultraviolet microscope that we term fibre optic microscopy with ultraviolet surface excitation (FUSE), using optical fibres for enhanced illumination control, compactness, and importantly, depth resolution(*13*). FUSE produced unique microscopic views of colonic polyp samples in real-time, enabling a new class of high-resolution mucosal imaging that we term surface histology. We showed that surface histology is widely interpretable after minimal training, by endoscopists and pathologists of varying experience. As an important step towards rapid widespread adoption, we also found that pathology foundation AI models unfamiliar with slide-free imaging were able to achieve near-human expert interpretation of FUSE after exposure to the same limited dataset used by human readers. FUSE enables a pathology-empowered ‘resect, diagnose, then discard’ approach to the endoscopy suite that promises to reduce polyp burden and enhance cost-effectiveness.

## RESULTS

### Clinical study of FUSE microscopy

Our group has developed FUSE microscopy, with a unique fibre optic approach for the oblique delivery of ultraviolet-C (also known as deep UV) light on the tissue sample (Fig. 1A). Our previous work(*13*) showed that optical fibres were capable of achieving a thin optical section from the sample surface, producing high quality images at magnifications from 4X to 50X, on fresh and fixed samples across a broad range of murine organs and fluorescence assays. For diagnostic applications, samples were stained with Hoechst for nuclear labeling and rhodamine for cytoplasm to mimic the traditional hematoxylin and eosin (H&E) specificity. For purposes of practical deployment in a rugged clinical environment, we selected a single magnification of 10X (measured lateral resolution 0.87 µm, field of view 750 µm x 560 µm), upgraded the platform with a custom high-speed stage (max speed 15 mm/s, max acceleration 80 mm/s^-2^) and a splash-proof 30 cm x 30 cm x 40 cm (height) enclosure. Polyps were resected, stained, and imaged on FUSE immediately after resection in a room adjacent to the endoscopy suite (Fig. 1B). Deployment involved a 2-person team and a collapsible table of size 1.2 m x 0.6 m for the inverted microscope and space for tissue preparation (Fig. 1C). The total time incurred from receiving a 1 mm x 1 mm fresh tissue sample, staining and mounting the sample and completing a scan over the full area with images available for review was about 2.5 minutes.

**Fig. 1.**
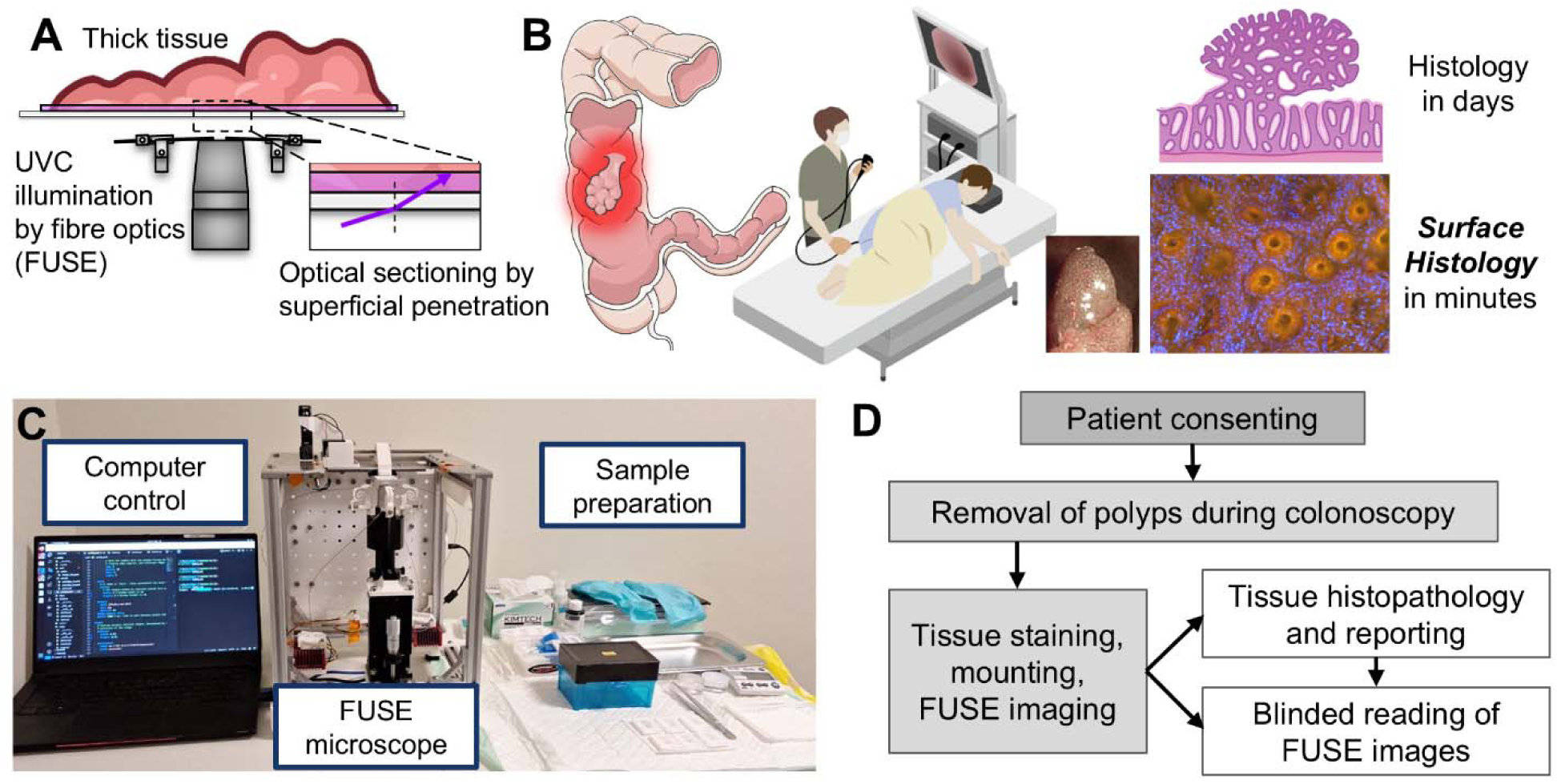
Fibre optic microscopy with ultraviolet surface excitation (FUSE) in colonoscopy: concept, implementation, and workflow. (A) FUSE uses optical fibres to illuminate a thick unsectioned sample with UVC light for enhanced depth resolution. (B) Endoscopists prefer relying on resource-intensive pathologist reporting for polyp diagnosis even though the real-time surface pattern can be revealing. FUSE enables a new intraprocedural readout from the surface pattern that we call ‘surface histology’, an intersection familiar to both specialties. (C) FUSE is space-efficient and can be deployed in close proximity to the procedure. Microscope has side covers removed to show internals. (D) Workflow of FUSE pilot study comparing blinded readings of FUSE images to gold-standard histopathology reports.

The study was performed in the Division of Colorectal Surgery, National University Hospital, Singapore from January to October 2024. The inclusion criteria were patients scheduled for screening colonoscopy and previously positive on a fecal (occult blood or immunochemical) test. 56 patients were recruited, from whom 28 yielded data for the study, and 79 polyps were imaged, from which 42 yielded usable datasets for the study (Table 1). After FUSE imaging, samples were sent for standard downstream histopathological processing, which provided gold-standard interpretations for the FUSE reading study (Fig. 1D).

**Table 1:**
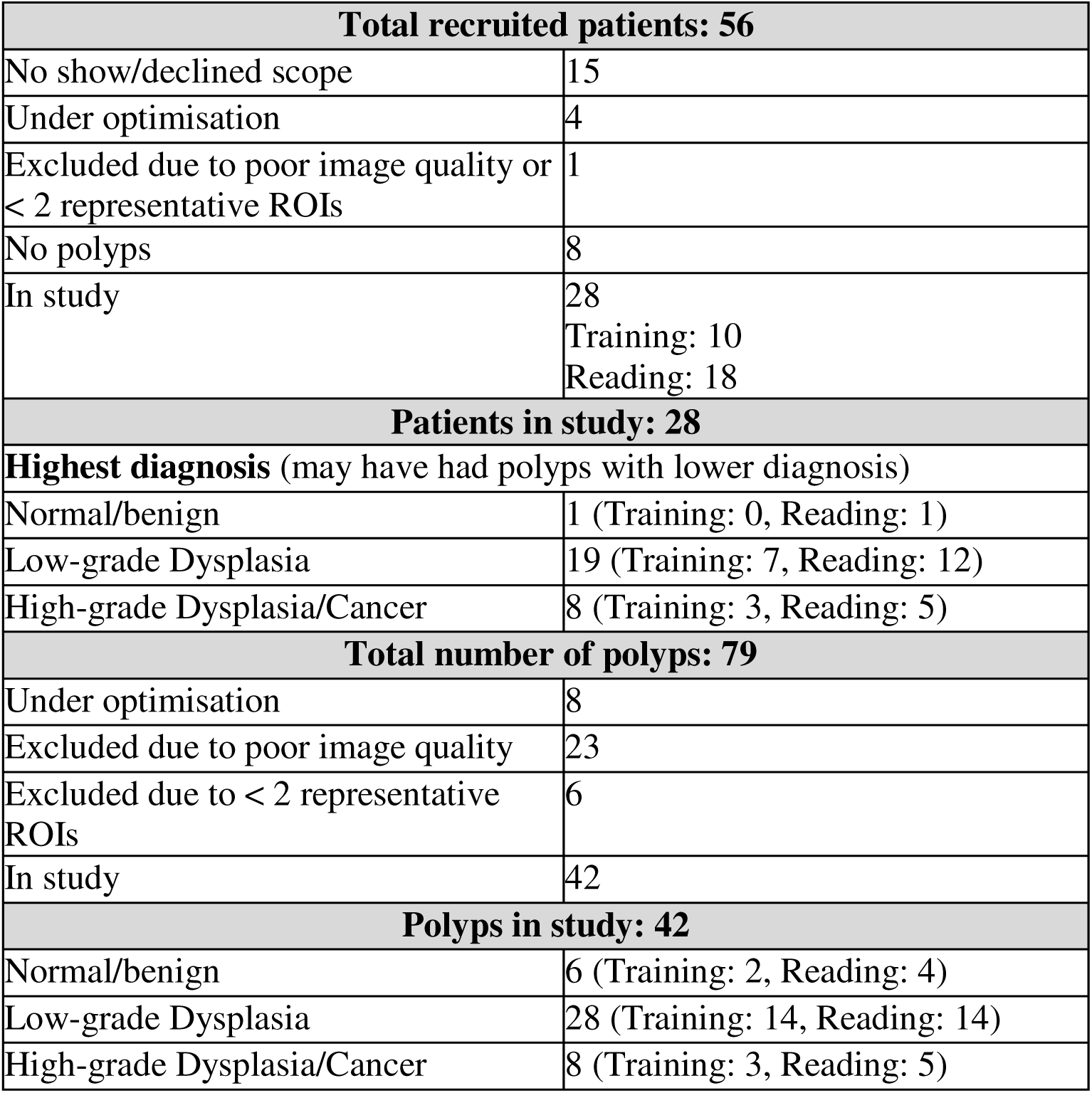
Summary of patients and polyps in study.

| **Total recruited patients: 56** |  |
| --- | --- |
| No show/declined scope | 15 |
| Under optimisation | 4 |
| Excluded due to poor image quality or < 2 representative ROIs | 1 |
| No polyps | 8 |
| In study | 28<br>Training: 10<br>Reading: 18 |
| **Patients in study: 28** |  |
| **Highest diagnosis** (may have had polyps with lower diagnosis) |  |
| Normal/benign | 1 (Training: 0, Reading: 1) |
| Low-grade Dysplasia | 19 (Training: 7, Reading: 12) |
| High-grade Dysplasia/Cancer | 8 (Training: 3, Reading: 5) |
| **Total number of polyps: 79** |  |
| Under optimisation | 8 |
| Excluded due to poor image quality | 23 |
| Excluded due to < 2 representative ROIs | 6 |
| In study | 42 |
| **Polyps in study: 42** |  |
| Normal/benign | 6 (Training: 2, Reading: 4) |
| Low-grade Dysplasia | 28 (Training: 14, Reading: 14) |
| High-grade Dysplasia/Cancer | 8 (Training: 3, Reading: 5) |

### Surface histology is reminiscent of both endoscopy and pathology

The gastrointestinal mucosa manifests patterns on the surface that have been widely studied as an important diagnostic signature of neoplasia(*14*). These mucosal patterns may be examined live with endoscopic narrow band imaging (NBI) or the equivalent blue light imaging (BLI), where descriptors for surface and vascular patterns such as those proposed by the Japan NBI Expert Team (JNET)(*15*) have revolutionized *in vivo* diagnosis. However, these images are generally obtained at a magnification and resolution far lower than microscopic histology, and are not expected to yield confirmatory pathological status.

To bridge the persistent gap between surface endoscopy and conventional histopathology, we propose the concept of ‘surface histology’, a histology-like microscopic view of the mucosal surface. The <300 nm ultraviolet (UV-C) illumination used in FUSE produces an optical section from the sample surface, hence FUSE is particularly suited for high-resolution mucosal surface imaging. We used FUSE to examine the intact mucosal surface of polyps without cutting into them, enabling a unique imaging modality that captures distinctive elements of endoscopy and pathology, presenting a fusion of traditionally orthogonal perspectives. Compared to NBI/BLI, FUSE is *ex vivo* but presents much higher sub-micron resolution views with nuclear contrast. Compared to histology, FUSE is lower resolution and views a non-standard surface plane, but yields pathological insight with far less processing and time. Hence FUSE does not replace but complements these techniques, and carves out a completely new niche in the colonoscopy paradigm.

Surface histology on resected polyps presents a similar imaging plane as endoscopy, but at sub-micron resolution and with multi-colour fluorescent stains mirroring the nuclear and cytoplasmic specificity of H&E. Normal colon mucosa (confirmed by pathology) is presented in Fig. 2A-E. The round crypts/pits were observed on endoscopy and recapitulated on FUSE at much higher magnification. The nuclear contrast of FUSE contributed powerful histopathology readouts unavailable in endoscopic practice such as nuclear size, density and polarity, and goblet cells. Virtually all prior work in slide-free microscopy have endeavored to recolor their fluorescence images to the purple and pink of H&E that pathologists are most familiar with(*5*, *8*, *16*, *17*). In this work, we deliberately retained the native colour of the images that arose from the fluorescence emission of the fluorophores, with a black/dark background typical in fluorescence microscopy and endoscopy. The decision came out of feedback from clinicians, including pathologists, who had commented that the native appearance of FUSE may have higher contrast than recoloured H&E. Also, we wished to avoid overly biasing the data appearance to a pathologist audience, with the intention of preserving cross-interpretability. As discussed in a later section, we recoloured the images only when presenting them to a pathology foundation AI model.

**Fig. 2.**
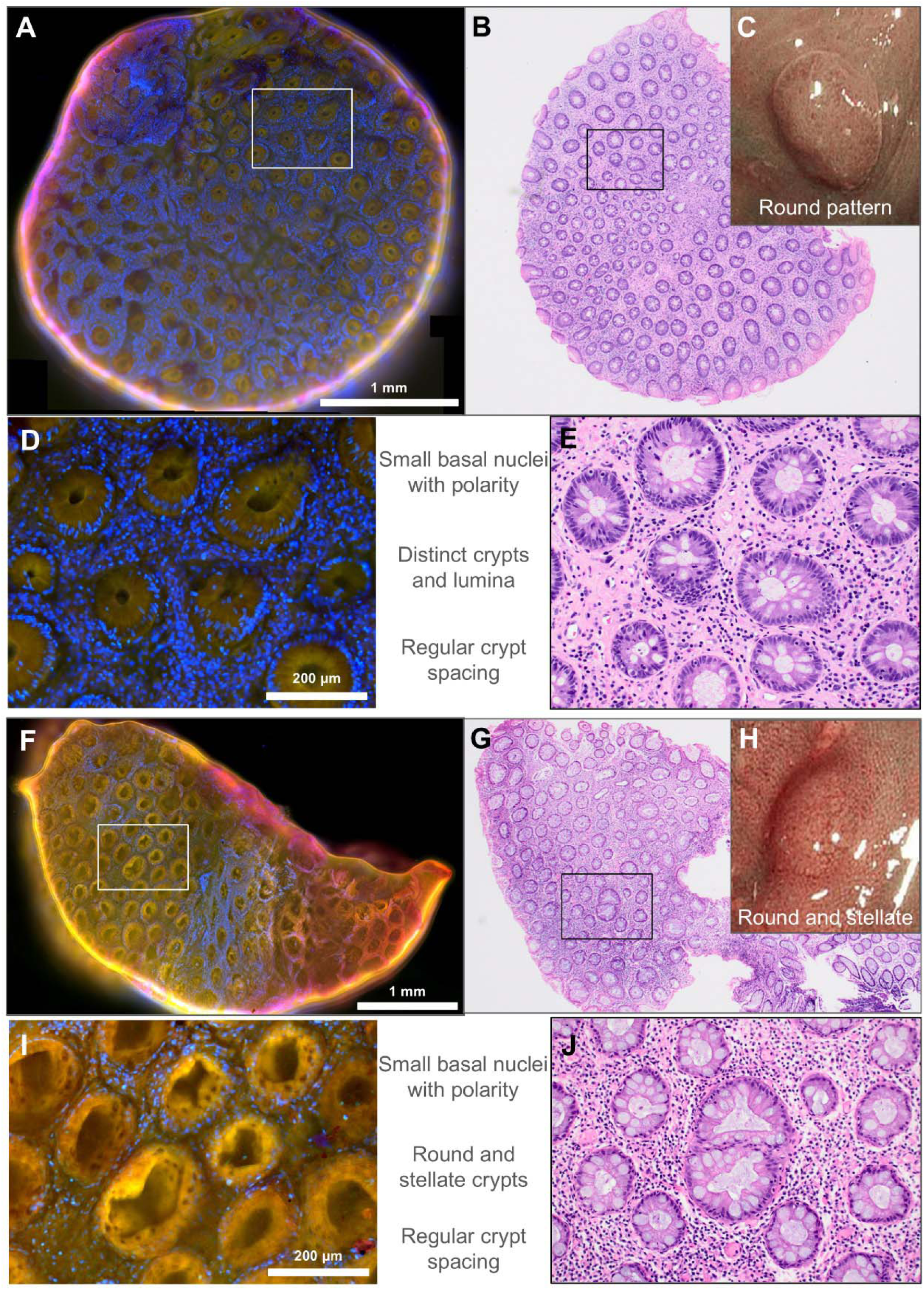
Exemplary FUSE surface histology of freshly excised, unsectioned (A-E) normal colonic mucosa and (F-J) benign hyperplasia show round and regular crypt organization similar to endoscopic pit pattern and histopathology. (A) FUSE imaging and (B) corresponding H&E histology and (C) endoscopic view of normal colonic mucosa, with accompanying insets (D-E) with suggested descriptors of nuclear size and polarity, crypt size, shape, and spacing applicable to both conventional histopathology and FUSE. (F-H) FUSE, H&E histology and endoscopic view of benign hyperplasia, with corresponding insets (I-J). Benign hyperplasia shows subtle changes in crypt architecture, which may include stellate crypts.

### Surface histology reveals signatures of polyp type and diagnosis

For major polyp types, FUSE captured the same suggestive mucosal pattern as endoscopy while reflecting confirmatory microscopic features that would normally require histological processing. Benign hyperplasia (Fig. 2F-J) showed mostly normal crypt and nuclear features with the additional characteristic star-shaped (‘stellate’) crypts on both endoscopy and FUSE. The ability to distinguish crypts with serrated features suggests that FUSE may also be able to identify the family of serrated polyp histologies. Compared to benign tissue, nuclear and crypt differences of polyps with low-grade dysplasia were subtle but qualitatively discernible. In addition, FUSE captured variations in crypt morphology occurring in low-grade dysplasia, such that tubular and tubulovillous adenomas could be clearly distinguished (Fig. 3A-J).

**Fig. 3.**
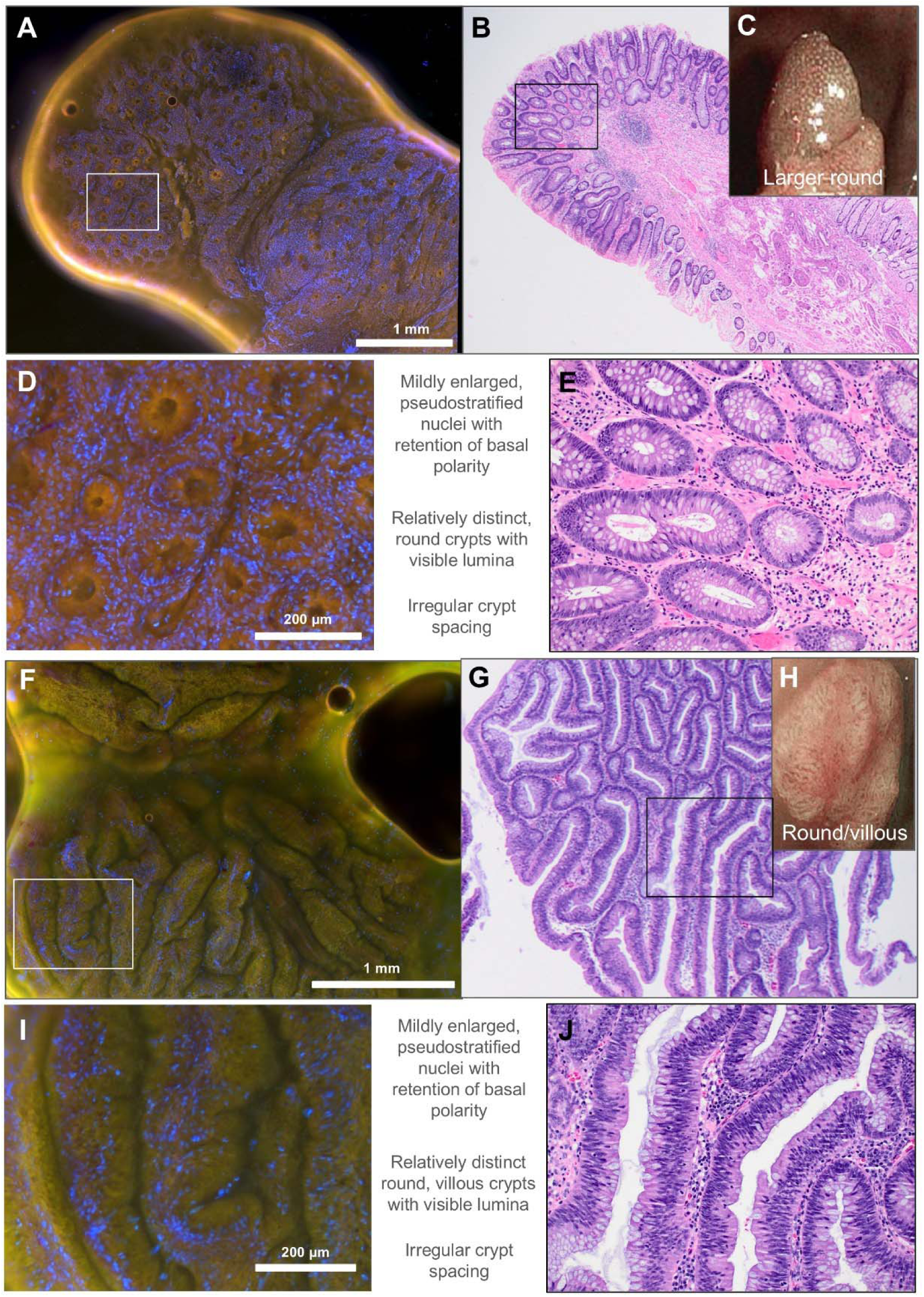
FUSE surface histology distinguishes low-grade dysplasia polyps (A-E) tubular adenoma and (F-J) tubulovillous adenoma. (A) FUSE imaging and (B) corresponding H&E histology and (C) endoscopic view of tubular adenoma, with accompanying insets (D-E) showing mild nuclear atypia and crypt irregularity. (F-H) FUSE, H&E histology and endoscopic view of tubulovillous adenoma, with corresponding insets (I-J) showing villous crypts. The microscopic resolution of FUSE over a large field of view enables not only clear determination of individual crypt shape but also an assessment of the villous component over a large area. Villous differentiation and consequently polyp type is an important indicator of cancer risk and a standard histopathology readout.

Samples of advanced neoplasia comprising both high grade dysplasia and cancer showed clear hallmarks analogous to known markers in histopathology and endoscopy, such as an irregularity or loss of crypt pattern (Fig. 4A-J). Nuclear features are a critical readout in pathology practice and could also be rapidly assessed through FUSE. Advanced neoplasia was strongly associated with nuclear proliferation and enlargement as reflected by density and size, analogous to the ‘nuclear-cytoplasmic ratio’ metric used by pathologists. While the endoscopic appearance (Fig. 4C, H) might be sufficiently indicative of advanced neoplasia to an experienced endoscopist, FUSE enabled a multidimensional interpretation – crypt visualization provided structural concordance with the endoscopic view, and fluorescence contrast contributed cellular detail akin to histopathology. Taken together, FUSE not only validates endoscopic findings but also enables a completely different perspective previously unavailable in the endoscopic suite. Notably, FUSE does not assess deeper or transected planes as is typical in regular pathology practice, and thus cannot determine depth of invasion, which would be required for definitive cancer diagnosis or staging.

**Fig. 4.**
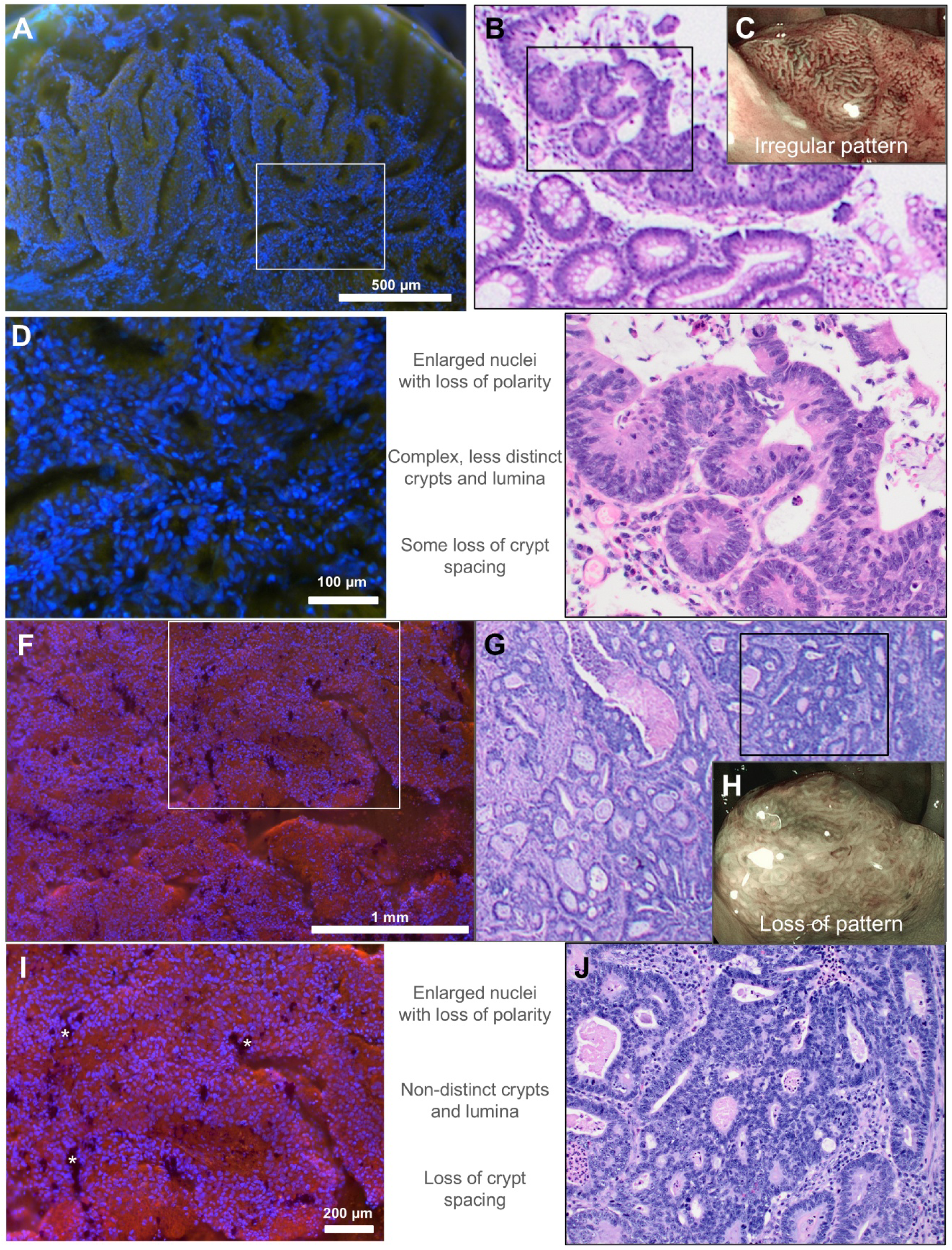
Nuclear contrast is a unique intraprocedural readout during endoscopy and an important marker of advanced neoplasia. (A) FUSE imaging and (B) corresponding H&E histology and (C) endoscopic view of high-grade dysplasia polyp, with accompanying insets (D-E) showing significant nuclear atypia and crypt irregularity. (F-H) FUSE, H&E histology and endoscopic view of cancerous sample, with corresponding insets (I-J) showing loss of crypt pattern, including an exemplary type of image artefact (*) on FUSE caused by stain precipitate that readers were trained to neglect. While endoscopic imaging in experienced hands already highlights changes in crypt architecture, the availability of nuclear contrast during procedures adds a new dimension of interpretation to the endoscopist, is straightforward to assess even at low magnification, and helps to align endoscopic assessment with histopathology practice.

### Blinded reading by human experts

Given the similarities of FUSE imaging with both mucosal patterns and polyp histology, we conceived that FUSE could be readily interpretable by both endoscopists and pathologists, making it the first ‘cross-interpretable’ endoscopic modality to our knowledge. To verify this hypothesis, we performed blinded reading studies individually with 1 non-clinician scientist (baseline) and 4 clinicians - a colorectal surgery consultant (qualified specialist) and resident (specialist in training), and a pathology consultant and resident. The study involved a brief training session of 19 polyps, and a reading test of 23 polyps in which readers were asked to review a set of FUSE images each from a different polyp (Supplementary Fig. 1), and predict their histopathology status based on 3 classes (normal/benign - 1, low-grade dysplasia - 2, and high-grade dysplasia or cancer – 3) while blinded to their histopathology reports. Readers also indicated their confidence level (high/low) when interpreting each image. There was no overlap of polyps or patients between training and test sets. In a test set of 4 normal/benign, 14 LGD and 5 HGD/cancer samples, the surgeon consultant and resident, and pathology consultant and resident achieved 3-class accuracies of 82.6%, 87%, 91.3%, and 91.3% respectively, with accuracies mostly higher among high-confidence reads (Table 2). Notably, when assessing test performance for the identification of high/low risk (class 3 versus class 1/2), all readers achieved perfect scores (100% sensitivity and specificity).

**Table 2:** Blinded readings by clinicians of surgical and pathology specialties and different training stages, and by pathology foundation AI models. A non-clinician scientist and a computed estimate of nuclear-cytoplasmic ratio served as human and computational baselines respectively for comparison. Both human and AI readers used the same training and test datasets, with virtual H&E recolouring performed only for AI readers. AI readers used a linear probe (3-class logistic regression) for classification. AI readers struggled to differentiate benign from low-grade dysplasia, but performed at human expert level for identification of advanced neoplasia (class 3).

| Reader | 3-class reading |  |  |  |  |  | Cls. 1+2 vs. Cls. 3 |  |
| --- | --- | --- | --- | --- | --- | --- | --- | --- |
|  | Normal/<br>Benign | Low Gr.<br>Dysplasia | High Gr.<br>Dysplasia<br>/Cancer | **Total**<br>**acc.**<br>**(%)** | High-<br>confi<br>reads | **High-**<br>**confi**<br>**acc. (%)** | **Sens**<br>**(%)** | **Spec**<br>**(%)** |
| Scientist | 2/4 | 14/14 | 4/5 | 87.0 | 11/23 | 100 | 80 | 100 |
| Colorectal Surgeon | 2/4 | 12/14 | 5/5 | 82.6 | 14/23 | 85.7 | 100 | 100 |
| Surgery Resident | 3/4 | 12/14 | 5/5 | 87.0 | 15/23 | 93.3 | 100 | 100 |
| Pathologist | 2/4 | 14/14 | 5/5 | 91.3 | 18/23 | 94.4 | 100 | 100 |
| Pathology Resident | 3/4 | 13/14 | 5/5 | 91.3 | 19/23 | 89.5 | 100 | 100 |
| N/C ratio | 0/4 | 14/14 | 1/5 | 65.2 | N/A |  | 20 | 100 |
| UNI2 | 0/4 | 13/14 | 4/5 | 73.9 | N/A |  | 80 | 94 |
| Phikon-v2 | 0/4 | 13/14 | 4/5 | 73.9 | N/A |  | 80 | 94 |
| Virchow2 | 1/4 | 14/14 | 5/5 | 87.0 | N/A |  | 100 | 100 |

### Foundation AI model interpretation

Here we explored the recent advent of pathology foundation AI models (FM), trained on massive amounts of histopathology whole slide image (WSI) scans. Despite the significantly lower image quality and resolution of slide-free FUSE compared to standard WSI, we hypothesized that the similarity of FUSE to histopathology could be exploited such that foundation models may make some sense of FUSE and achieve few-shot learning on a relatively small dataset. We used three popular open-weights pathology models UNI2(*18*), Phikon-v2(*19*) and Virchow2(*20*) to obtain feature embeddings for the same FUSE images from the blinded reading study, only recoloured to virtual H&E (Supplementary Fig. 2) as previously reported by others(*17*). As a computational baseline we calculated a set of 3 features based on simple estimates of the nuclear/cytoplasmic ratio (area of Hoechst signal divided by area of rhodamine signal), a commonly used metric by pathologists and a straightforward indicator of malignancy. The features from the same training set used by the human readers were used to train a multi-class logistic regression, a deliberately simple model (also known as a linear probe) intended to test the strength of the features. Using the same test set (unseen in training) as the human readers, all three FMs achieved 3-class accuracy and 2-class diagnostic performance comparable to humans, with Virchow2 performing the best (87% 3-class accuracy, 100% sensitivity and specificity to high/low risk) (Table 2).

## DISCUSSION

Endoscopy is routinely used in the examination of virtually all luminal organs in the body, and the ability of the endoscopist to rapidly assess any anomalous findings and decide next steps is critically important to patient care. It has long been suggested that endoscopists may derive histological clues by close study of the luminal surface pattern, and the assessment of colorectal polyps has been a touchstone of this paradigm. Herein we propose that the surface pattern becomes an even more powerful predictor after resection, by producing a surface histology readout via slide-free FUSE microscopy that harmonizes the diagnostic language of both endoscopy and pathology for accurate polyp assessment in real time. Improving the efficiency of luminal tissue diagnosis during endoscopic procedures through a new modality that is cross-interpretable could be an important milestone for the real-time histological determination of resected tissue, providing much needed support for endoscopist decision-making and promising to evolve the paradigm into what may become known as ‘resect, diagnose, then discard’. As complex therapeutic procedures such as endoscopic mucosal resections and submucosal dissections become more widely performed alongside a risk of residual/recurrent disease after excision(*21*, *22*), margin assessment in endoscopy analogous to surgery could be an emerging need, potentially requiring real-time histopathological determination akin to frozen sections.

The pursuit of purely endoscopic diagnosis via a myriad of optical non-excisionary approaches including optical coherence tomography(*23*, *24*), confocal endomicroscopy(*25*), Raman spectroscopy(*26*), autofluorescence(*27*) and others continues to be an important direction of research and development. It is also conceivable that FUSE and surface histology could be eventually extended to a fully endoscopic *in vivo* approach, notwithstanding ongoing studies into the safety of brief UV-C exposures on living cells and tissue(*28*). It is worth noting that extremely high “1000x” magnification endoscopes with cellular resolution have been commercially available for years(*29*) but have never achieved mainstream appeal. The technical challenge of capturing stable images *in vivo* at high magnifications as well as their esoteric interpretation, the lack of nuclear specificity, and ultimately the high likelihood of polyps being resected regardless of endoscopic appearance may explain the limited impact of *in vivo* diagnosis. The resection of polyps and their intraprocedural microscopic examination may be arguably one of the quickest paths towards reducing polyp diagnostic burden by uniquely engaging with pathology.

Intraoperative imaging and microscopy has been a longstanding effort by bioengineers, but the continued necessity for clinicians to ultimately interpret and sign-off these new unfamiliar modalities dampens claims of improved efficiency and cost savings, and is a thorny issue in practical non-research scenarios. Even as visual artificial intelligence (AI) has revolutionized medical imaging interpretation, conventional approaches still demand the collection of a large amount of data with the new modality correlated with gold standard labels for supervised learning, with concerns of generalizability of this small training distribution unresolved. The applicability of pathology FMs to not only histopathology WSIs but also slide-free fluorescence microscopy is an important finding that greatly expedites the translation of intra-operative imaging, by relaxing the necessity for either extensive clinician retraining or large bespoke datasets for customized AI. Relieving the endoscopist from the burden of additional real-time image interpretation further lowers the barrier to entry. There is an assumed caveat that the novel modality should sufficiently approximate histology for such FMs to be applicable, although the extent of similarity to the FM’s vast training set that is necessary remains unclear and is an interesting direction of future investigation.

This pilot study’s implications are limited by a small enrollment at a single site and a significant fraction of data being excluded, resulting in just 42 unique polyps (including 8 samples with advanced disease) out of 79 being analysed. The study also lacked data and validation of serrated polyps, an important subset of histologies that would likely arise in a future prospective recruitment. Pilot studies usually have modest enrolments(*30*) and may be seen as a precursor to a future clinical trial. The moderate usable yield (42/79) of imaged polyps was due to the presence of image artefacts or poor quality images, and would need to be substantially improved before practical implementation in the clinic. Attempting to flatten a fresh and/or irregularly shaped sample for optimal imaging coverage sometimes led to distortion of delicate tissue architecture, which affected image quality. In a practical scenario, an occasional polyp that yields inadequate quality FUSE data could simply be sent to pathology for avoidance of doubt, and should not diminish the added diagnostic value that FUSE may offer on the majority of polyps. Further study into the FUSE image artefacts that may occur, protocol enhancements to reduce artefacts and improve quality consistency, as well as enhancing the robustness of human and AI readers to suboptimal imaging would be needed to achieve real-world translation to a broader range of settings and practitioners.

## MATERIALS AND METHODS

### Study design

The inclusion criteria were patients with a fecal immunochemical or occult blood test positive result and scheduled for colonoscopy. 56 patients who were fecal immunochemical test-positive were recruited from the Division of Colorectal Surgery, National University Hospital, Singapore. 15 patients were later excluded due to scheduling constraints or withdrawal from study, 8 did not have any polyps, 4 were used for optimisation and 1 had poor image quality, leaving 28 patients whose data were used for reading (Table 1). Written informed consent was obtained from all subjects. Research was performed with approval from the National University Hospital Domain Specific Review Board (DSRB).

### Sample preparation and imaging

Polyps were imaged immediately after resection, with the FUSE microscope deployed in a room adjacent to the endoscopy suite, a setup inspired by frozen section rooms in close proximity to the surgical procedure in some centres. Resected polyps were processed individually. Each polyp was first rinsed with deionised water, then placed in a mixture of 0.2 mg/mL Hoechst 33342 and 1mg/mL Rhodamine B. After 1 minute, the polyp was removed from the staining mixture and rinsed in deionised water. Hoechst enabled nuclear visualisation and Rhodamine B provided cytoplasmic contrast (fluorescent analogues to H&E). The rinsed polyp was dried before being placed onto a custom imaging cassette for imaging. After imaging, samples were sent for conventional histopathology to obtain gold standard pathologist interpretation. The FUSE microscope was previously described(*13*). In brief, the microscope used an inverted widefield design. Four optical fibres (57-070, Edmund Optics) delivered deep ultraviolet (DUV) illumination from two LEDs (ILR-OV01-O275-LS030-SC201, Osram) to the sample. A 10X 0.25 NA objective (43-907, Edmund Optics) was used as the lens. A colour camera (BFS-U3-120S4C-CS, Teledyne FLIR) was used to capture images.

### Reading data preparation

Pathologists evaluated routine H&E-stained formalin fixed paraffin embedded (FFPE) histology slides of the polyps hours to days after colonoscopy procedure and FUSE imaging, and provided deidentified patient diagnosis for each excised polyp. Out of 79 polyps imaged, 8 polyps were used for initial protocol optimisation and did not yield usable data, and 23 polyps had inadequate image quality for reasons such as out-of-focus, inadequate nuclear staining, or excessive artefacts. We further imposed an exclusion criterion for samples that did not yield at least two infocus FOVs displaying representative diagnostic features, to ensure rigor of feature identification and robustness of reading; 6 polyps were excluded. This resulted in 42 polyps used in the reading study, a yield of 53% (42/79). Patients and polyps were grouped by their pathological diagnosis into 3 classes (Normal/Benign, Low Grade Dysplasia and High Grade Dysplasia or Cancer), and subsequently allocated into training and reading datasets (Table 1).

Training and reading datasets were prepared for the study reading. The training dataset was designed to include representative images and annotated diagnostic features of a total of 19 polyps from 3 classes (2 polyps from Normal/Benign, 14 polyps from Low Grade Dysplasia and 3 polyps from High Grade Dysplasia or Cancer). One of the polyps had 2 regions of interest shown in training, hence a total of 20 images were shown from 19 polyps. Training was performed to adequately train readers to identify features (nuclear and crypt) that would aid classification, and to disregard artefacts. The reading dataset consisted of images from each of 23 polyps from 3 categories (4 polyps from Normal/Benign, 14 polyps from Low Grade Dysplasia and 5 polyps from High Grade Dysplasia or Cancer). Readers were trained with the training dataset preceding the blind reading test, and were able to reference the training material during the reading. Readers assigned diagnoses of Class 1, 2, or 3, together with their confidence level (high/low confidence) to each FUSE image from the reading dataset.

### Study readers

Blinded FUSE image reading was performed individually by a set of human experts comprising a consultant colorectal surgeon, a consultant pathologist with subspecialty in gastrointestinal pathology, a surgical resident, a pathology resident, and a non-clinician scientist. The consultant surgeon had performed the endoscopies and polyp resections in the study and the consultant pathologist had performed the corresponding histopathology readings as per standard clinical practice, but both had not previously reviewed FUSE data beyond a few anecdotal examples and had also undergone a >3 month washout period since their last look at any FUSE data or corresponding histology data. The surgical resident, pathology resident, and non-clinician scientist had not previously participated in the study and were unfamiliar with FUSE data.

Three open-weights foundation models (FMs) for pathology, namely UNI2(*18*), Phikon-v2(*19*) and Virchow2(*20*) were obtained and used to generate embeddings for FUSE images. The FMs used the same training set (20 images from 19 polyps) and test set (23 images from 23 polyps) as the human readers. The FUSE images in RGB were unmixed into 2-channel images with standard non-negative matrix factorization and virtually recoloured to H&E using a publicly available Python library(*17*). The FUSE images for the FMs were recoloured but not the images for human readers, because the FMs were known to be trained on H&E colours and the team deemed it reasonable for the FMs to see images of the same colour scheme. Since the FMs took square images as inputs, the 4000x3000 FUSE images were cropped to 3000x3000 and resized to 224x224 then given to the models for embedding generation. As a computational baseline, nuclear/cytoplasmic ratio was estimated by taking the positive area of the nuclear channel divided by the positive area of the cytoplasmic channel for a given field size, then taking the maximum obtained over the entire image. Three field sizes were used: 4000x3000 (entire image), 2000x1500, and 1000x750 to obtain a set of 3 features per image. The training set embeddings were used to train a logistic regression model for classification, known as a ‘linear probe’ to the machine learning community. The optimisation algorithm used was L-BFGS and the L2 regularization parameter λ was 100/(MC) where M was the embedding dimension and C=3 was the number of classes, as is standard practice. The logistic regression models were then used to produce predictions (Class 1, 2, or 3) on each image in the test (reading) set.

### Data analysis

Reading data for both human and AI readers were analysed in the same way. Predictions were checked against the histopathology gold standard, and accuracies per class were obtained. Accuracies for readings that were made with high confidence as indicated by the human reader were also calculated. To obtain sensitivities and specificities to advanced neoplasia (High Grade Dysplasia/Cancer), the Normal/Benign and Low Grade Dysplasia categories were pooled to produce a 2-class reading.

## Data Availability

All data needed to evaluate the conclusions in the paper are presented. Experimental data underlying the results, and code used for data analysis are available from the corresponding authors upon reasonable request.

## Acknowledgments

We thank Drs. Sunny Wong and Lucas Cahill for their feedback on the manuscript.

## Funding

Singapore National Research Foundation grant NRFF13-2021-0002 (KL) Agency for Science, Technology and Research grant M22K2c0089 (KL)

## Author contributions

Conceptualization and methodology: ASKY, NH, KHT, GSTS, DKHC, KL

Investigation and analysis: ASKY, NH, KHT, GM, RRYS, RMLL, OZJL, SST, GSTS, DKHC, KL

Supervision: DKHC, KL

Writing: ASKY, KHT, GM, RRYS, GSTS, DKHC, KL

## Competing interests

ASKY, GM and KL are inventors on patent filings related to FUSE technology. Other authors have no competing interests.

## Notes

### Funding Statement

This study was funding by the following grants.
Singapore National Research Foundation grant NRFF13-2021-0002 (KL)
Agency for Science, Technology and Research grant M22K2c0089 (KL)

### Author Declarations

Domain Specific Review Board (DSRB) of the National University Hospital (NUH) gave approval for this work

