## Supplementary Materials for "Slide-free surface histology enables rapid colonic polyp interpretation across specialties and foundation AI"

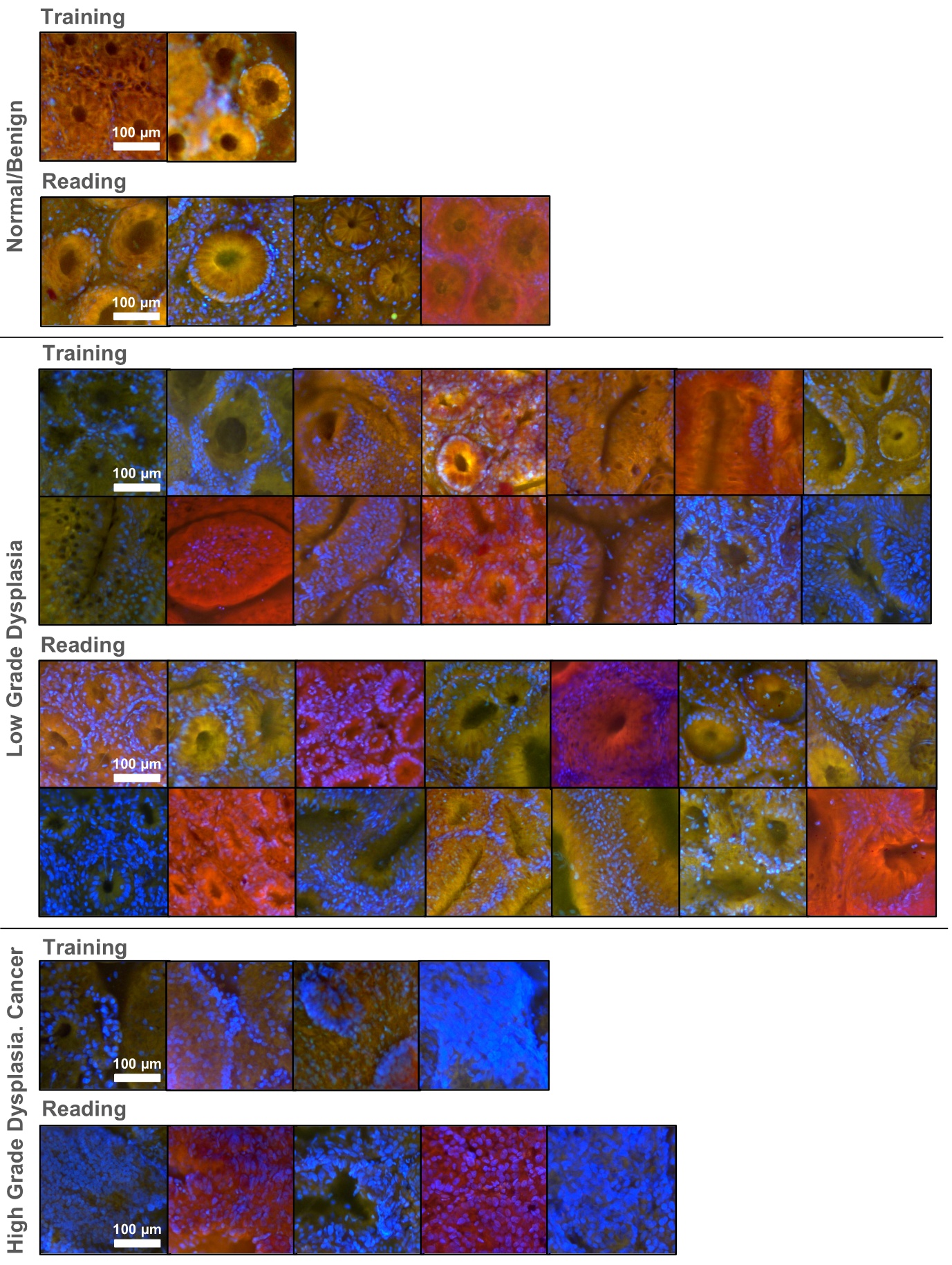

**Supplementary Fig. 1: Exemplary crops from each image used in the blinded reading study.** Cropped regions selected to illustrate distinctive features and wide variability in pattern and color. Actual images used were larger, covering one field of view (750 µm x 560 µm).

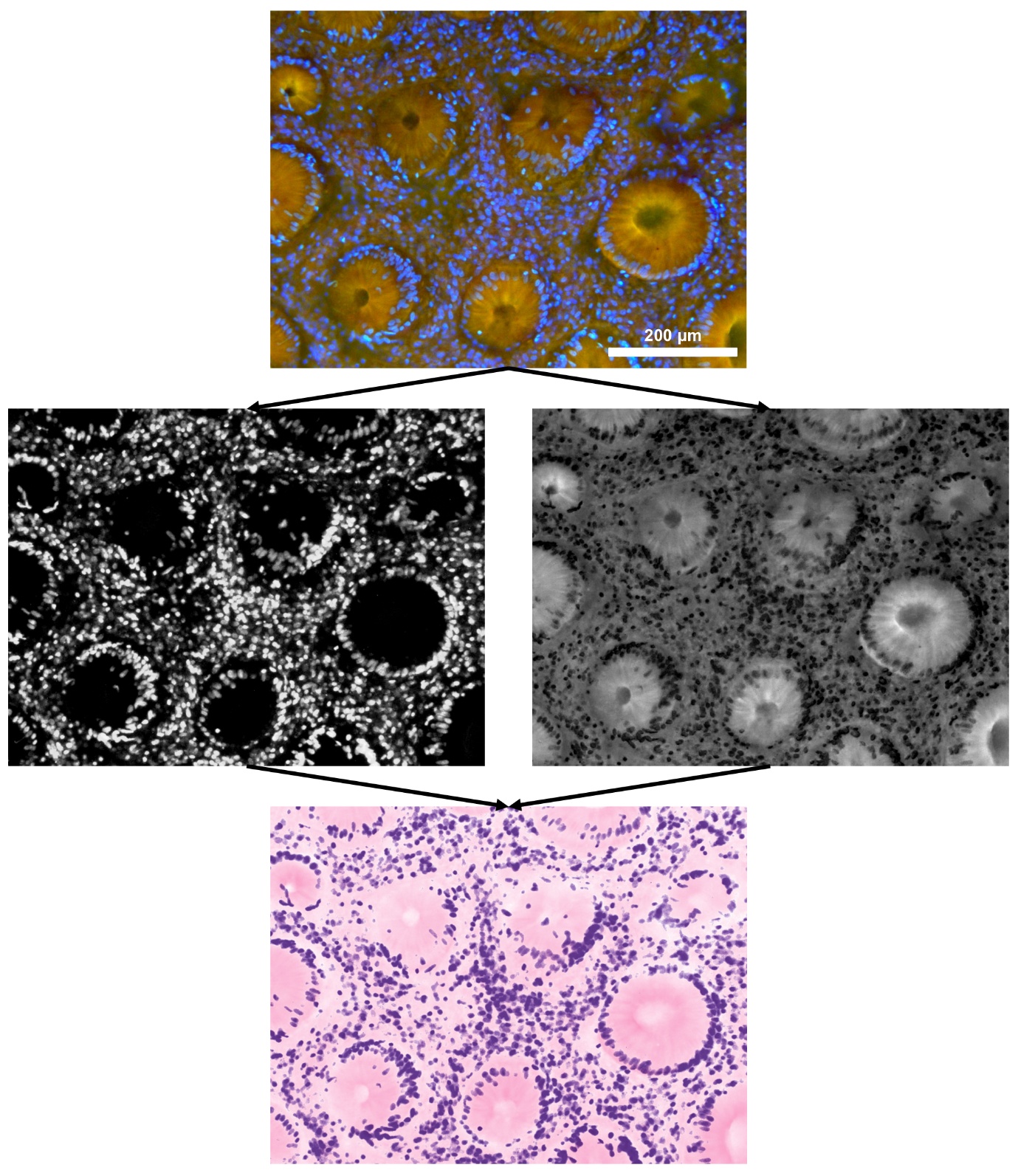

**Supplementary Fig. 2: Workflow of FUSE image recoloration to pseudo-H&E.** Colors were unmixed then recombined using a previously reported virtual transillumination algorithm.
